# Curvature-based 3D dental comparison to identify trauma-induced surface changes in human teeth: A forensic comparison study

**DOI:** 10.1101/2025.06.19.25329914

**Authors:** Anika Kofod Petersen, Rubens Spin-Neto, Palle Villesen, Line Staun Larsen

## Abstract

**Objectives:** This study evaluates the performance of a curvature-based 3D dental comparison method - the keypoint pipeline - for forensic identification, assessing the effects of standardised blunt force trauma on human dentitions.

**Methods:** The dental arches in ten human jaw specimens (five maxillae, five mandibulae) were scanned using two intraoral 3D scanners before and after exposure to controlled blunt trauma delivered via a drop tower mechanism applying approximately 3154 Newton of force. Trauma outcomes were documented through high-speed video, digital photography, and 3D scanning. Post-trauma scans were processed using the keypoint pipeline, which quantifies dental surface similarity by comparing curvature signatures. An all-vs-all comparison was conducted between pre- and post-trauma scans, including cross-scanner evaluations.

**Results:** Despite consistent trauma application, fracture patterns varied by jaw type, with mandibular fractures typically occurring in the frontal plane in the side segments and maxillary fractures in the sagittal plane in the midline suture. The keypoint pipeline successfully scored 92.5% of the true matches to be the best matching comparison, even in the presence of significant structural damage and tooth displacement. Matching pairs yielded lower dissimilarity scores (mean: 0.55) compared to mismatches (mean: 0.78), indicating that curvature features were sufficiently preserved post-trauma.

**Conclusions:** These findings support the integration of curvature-based 3D dental surface analysis into forensic odontology workflows, particularly in disaster victim identification scenarios involving blunt force trauma.

**Clinical Significance:** This study demonstrates that comparison of position invariant curvature keypoints might aid in *post mortem* identification cases where dentitions have been subjected to severe blunt force trauma.

**Highlights:**

- Blunt force trauma significantly alters the dentition surface
- Identification of blunt force trauma victims is difficult
- Comparison of placement-independent curvature keypoints might aid in trauma scenarios
- Curvature keypoints are resilient towards trauma conditions
- Surface keypoint comparison might help in the identification process after blunt force trauma

## INTRODUCTION

In the case of disasters, victims need to be identified. According to INTERPOL three primary identifiers are used for disaster victim identification, namely DNA, fingerprints and dental comparison [1–4]. For dental comparison, a forensic odontologist will compare the *post mortem* (PM) dentition with information from *ante mortem* (AM) dental records of possible victims to find identifying traits [1–6].

When comparing an AM dental record with a PM dentition, the forensic odontologist relies on congruence between the descriptions in the AM dental record and the PM dentition [1–6]. A description could be the placement of fillings or crowns, or agreement between AM and PM X-ray images [1–6]. In the absence of dental work, such congruence can be difficult to establish.

In recent years, efforts have been made to digitize parts of the dental comparison, by comparing AM 3D intraoral scans with PM 3D intraoral scans [7–18]. Such a comparison can potentially quantify dentition similarity, even when there is no dental work, giving the forensic odontologist an advantage when comparing dentitions. A quantitative similarity score can both be used to give an indication of identity, while it can also be used to sort the AM dental records for examination, especially in disasters with many victims [11,14,17]. Furthermore, such a method helps to address past criticism of the discipline as being excessively subjective.

So far, these 3D comparison studies have investigated various scenarios, including single, extracted teeth, heat exposure and digital partialisation [11,14,17].

In a disaster, the dentition might be subjected to blunt force trauma in various ways [19]. It could be impact from a traffic/high-speed incident, falling from great heights, explosions or other high-impact scenarios [19]. Even though the teeth are the hardest material in the human body and they are very resistant to many types of traumas, many trauma scenarios are still expected to have an impact on the dentition surface [6,19].

Dental blunt force trauma has been described in a forensic context, but to the best of our knowledge there has not been any study on the quantitative dental similarity of human dentitions before and after exposure to blunt force trauma [19,20]. After exposure to blunt force trauma, at least part of the dentition surface is expected to have changed. These changes could include, but are not limited to, infractions, enamel chipping off, fractured teeth, and missing teeth. Such changes to the dentition surface could potentially disturb automatic dental surface comparison to a great extent [11,14,17]. Previously the keypoint pipeline methodology for automatic 3D dental comparison within forensic odontology identification has been proposed [11,14,17]. But since this methodology relies on key surface curvatures to be present in both AM and PM data, blunt force trauma might constitute a problem for the methodology. But to what extent blunt force trauma might interfere with dental comparison using the keypoint pipeline has yet to be investigated.

This study aims at testing the previously developed keypoint pipeline method for quantitative dental comparison, specifically on human dentitions subjected to blunt force trauma [11,14,17]. This study therefore aims at testing the robustness of the keypoint pipeline for dental comparison in a context where the dentition surface is expected to have changed to some degree. Furthermore, the combination of both descriptive and quantitative comparison of human dentitions before and after blunt force trauma will add to the understanding of dental surface changes in blunt force trauma scenarios.

## MATERIALS AND METHODS

This *ex vivo* study was exempt from ethical approval (request 279/2017). The specimens were obtained from individuals who had donated their bodies to science; no information regarding the donors’ gender, age, or cause of death was available. All procedures were performed in accordance with the ethical standards of the institutional and/or national research committee and with the 1964 Helsinki Declaration and its later amendments or comparable ethical standards.

A total of 5 upper jaws and 5 lower jaws were scanned using two 3D intraoral scanners (PrimeScan AC and PrimeScan Connect, Dentsply-Sirona, Bensheim, Germany) before and after being subjected to standardised experimental blunt force trauma. The initial impact of the trauma was estimated to be 3,154 Newton, equal to approximately 321 kilograms of force (see supplementary material 1). Inspired by Houg et al. [20] the blunt force trauma setup was in the style of a drop tower, where a weight (2 kg) was being dropped from a fixed height (200 cm) directly onto the approximate occlusal plane of a fastened jaw, fastened to a rigid surface. Details of the trauma setup can be seen in Figure 1 and can be found in the experiment protocol in Supplementary Materials 1.

**Figure 1.**
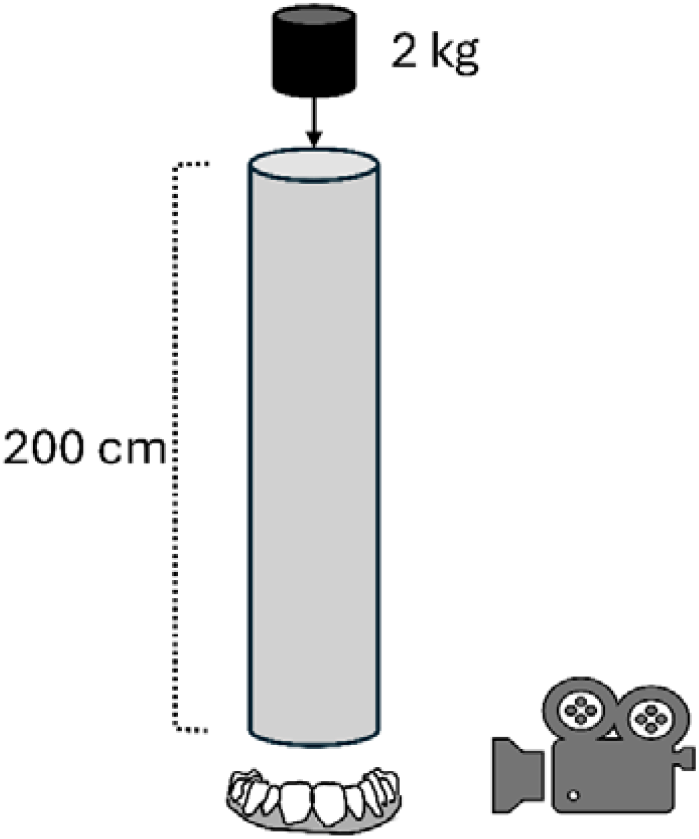
Experimental setup of the standardised blunt force trauma. A 200 cm drop was ensured by a pipe, guiding the falling object which weighed 2 kg. A high-speed camera recorded the impact to estimate the force of the impact.

The specimens were documented in the style of 3D scans and digital photos before and after the experiment, and the blunt force trauma was documented with high-speed camera footage during the impact.

The soft tissue part of the 3D scans was manually removed from all 3D scans, before subjecting them to keypoint detection and keypoint representation as described by the keypoint pipeline [11,14,17]. The keypoint pipeline is a previously proposed processing pipeline that compares curvature signatures on the dental surfaces between 3D dental scans to quantitatively evaluate similarity [11,14,17]. Since the position of some of the teeth changed during the trauma, the keypoint placement factor was disregarded from the scoring scheme when comparing dentitions [17]. This is not expected to change the performance of the keypoint pipeline scoring scheme, as the contribution of keypoint placement to the final score has been observed to be minimal, and the score without this factor has been reported to perform almost as well [17]. In this study, a score closer to zero would indicate a match, and a score closer to one would indicate a mismatch, as the scoring scheme indicates dissimilarity [14,17].

The comparisons were made in an all-vs-all manner, where all the scans before trauma were compared with all the scans after trauma, including comparisons between the two different scanners.

## RESULTS

Even though the trauma exposure was standardised by a drop tower, the trauma seen in the specimens varied significantly. Figure 2 shows the progression of the blunt force trauma exposure for one of the specimens.

**Figure 2.**
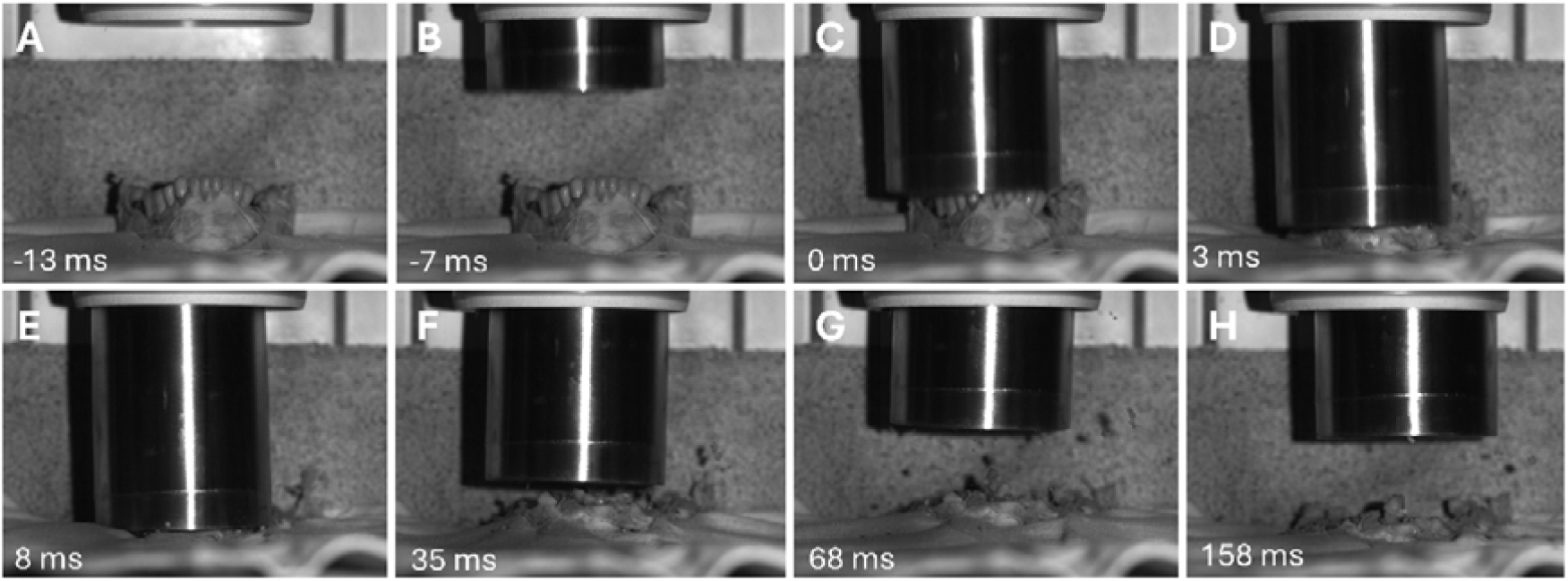
Still photographs of a mandibular specimen (ID05) during the experimental blunt force trauma. This specimen, in particular, suffered significant damage during the experiment. Time indicates milliseconds (ms) from first impact.

During the experiment, all 10 jaws suffered trauma to the bone and dental structures. For the lower jaws, fractures were mostly seen in the frontal plane, between canines and premolars, between premolars and molars (Figure 3), or between molars, while for the upper jaws’ fractures were observed in the sagittal plane in the midline suture (Figure 4). Tooth crown fractures were observed to varying degrees in both upper and lower jaw specimens (Figure 4 and 5). A single specimen appeared with comprehensive fixed prosthodontics and suffered multiple root fractures and displacement of all teeth. More teeth were seen to be intruded in the lower jaws (Figure 6) and displaced in the upper jaws. A tooth exarticulation occurred in two lower jaws (Figure 3), while more prosthetic work was separated from the tooth abutments/lost due to root fractures in the upper jaws.

**Figure 3.**
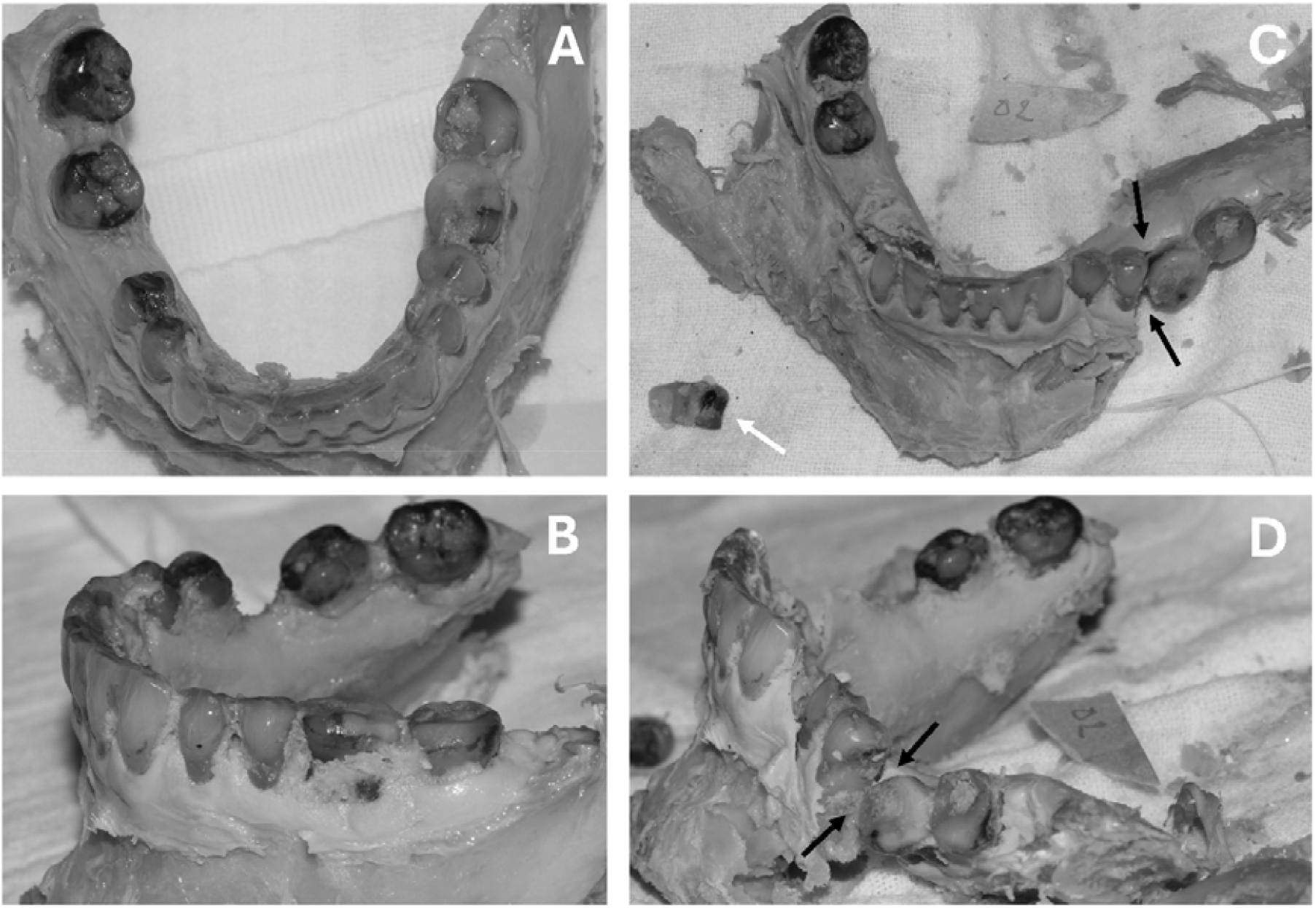
One of the mandibular specimens (ID02) before (A and B) and after the experimental trauma (C and D). The white arrow in C) points at tooth 45 that was exarticulated from the alvolar socket. A jaw fracture between tooth 35 and 36 displaced the posterior left part of the mandible (black arrows in C) and D)). The anterior segment was found intact.

**Figure 4.**
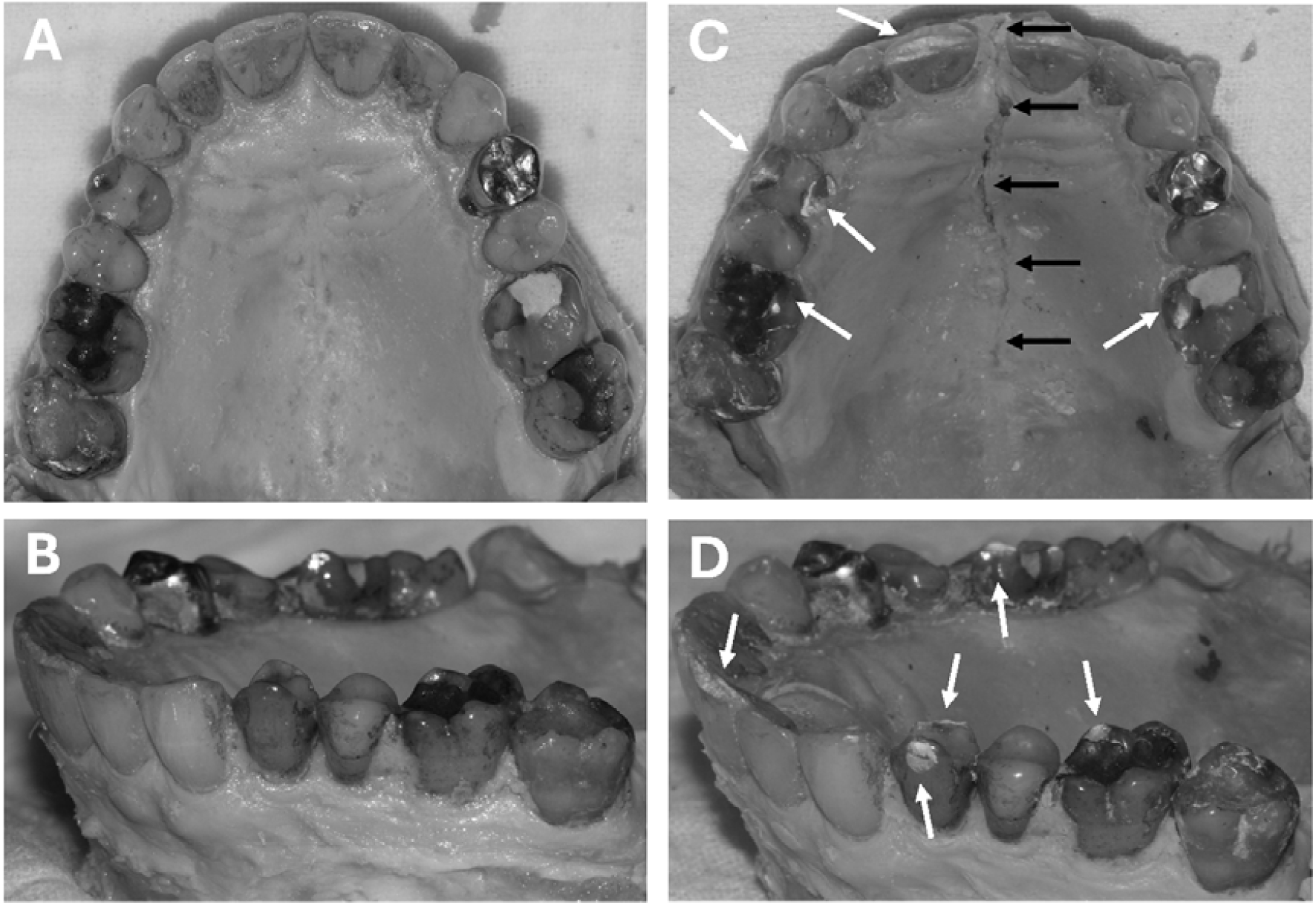
One of the maxillary specimens (ID10) before (A and B) and after the experimental trauma (C and D). The white arrows point at areas with obvious damage to the enamel (tooth 16, 14, 11 and 26). The black arrows mark a distinct fracture line along the maxillary midline suture (sutura palatina mediana).

**Figure 5.**
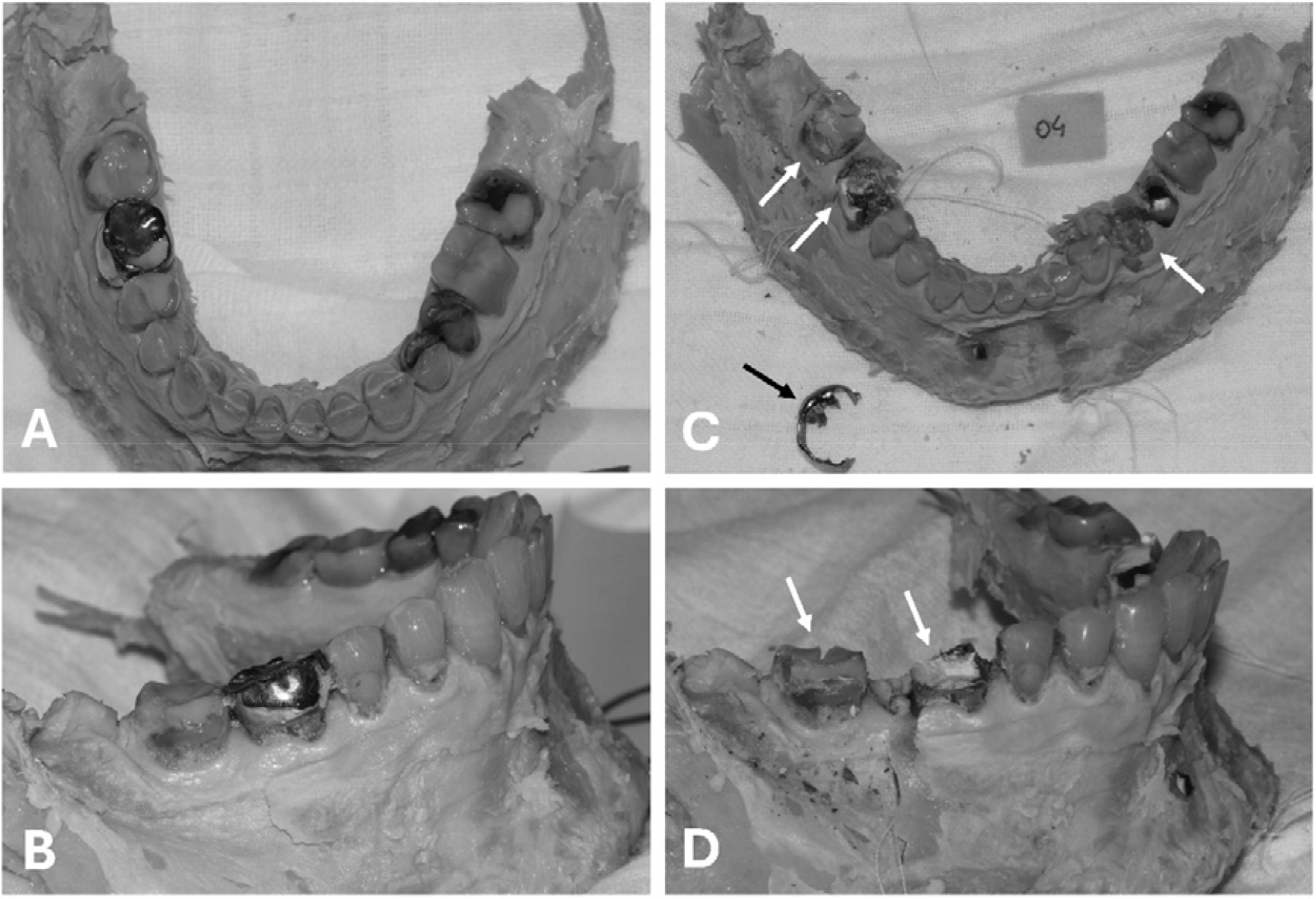
One of the mandibular specimens (ID04) before (A and B) and after the experimental trauma (C and D). The white arrows point at teeth where substantial parts of the tooth crown / restorative treatment suffered extensive damage during the test. In C) the prosthetic partial crown from tooth 46 can be seen broken next to the jaw (black arrow).

**Figure 6.**
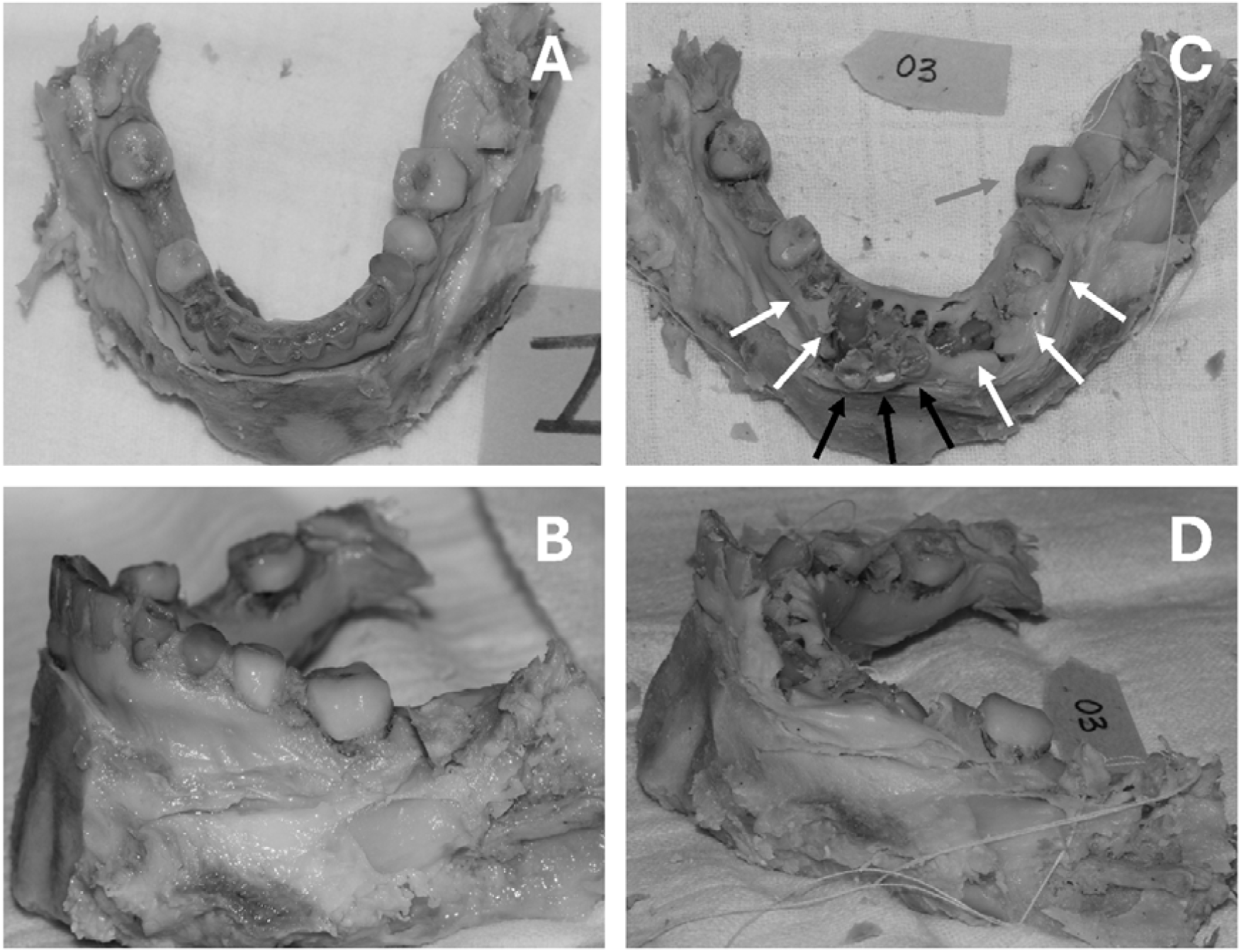
One of the mandibular specimens (ID03) before (A and B) and after the experimental trauma (C and D). The white arrows in C) point at tooth 44, 43, 33, 34 and 35 that were severely intruded into the bone. Teeth 42, 41 and 31 were displaced facially from their alveolar sockets (black arrows in C)) and 36 displaced lingually (grey arrow in C)).

When quantifying dentition surface similarity using the keypoint pipeline, the dissimilarity score was in most cases able to differentiate between matches and mismatches, despite the use of different scanners and the extensive destruction due to blunt force trauma. This is seen by the dark-colored diagonal in Figure 7.

**Figure 7.**
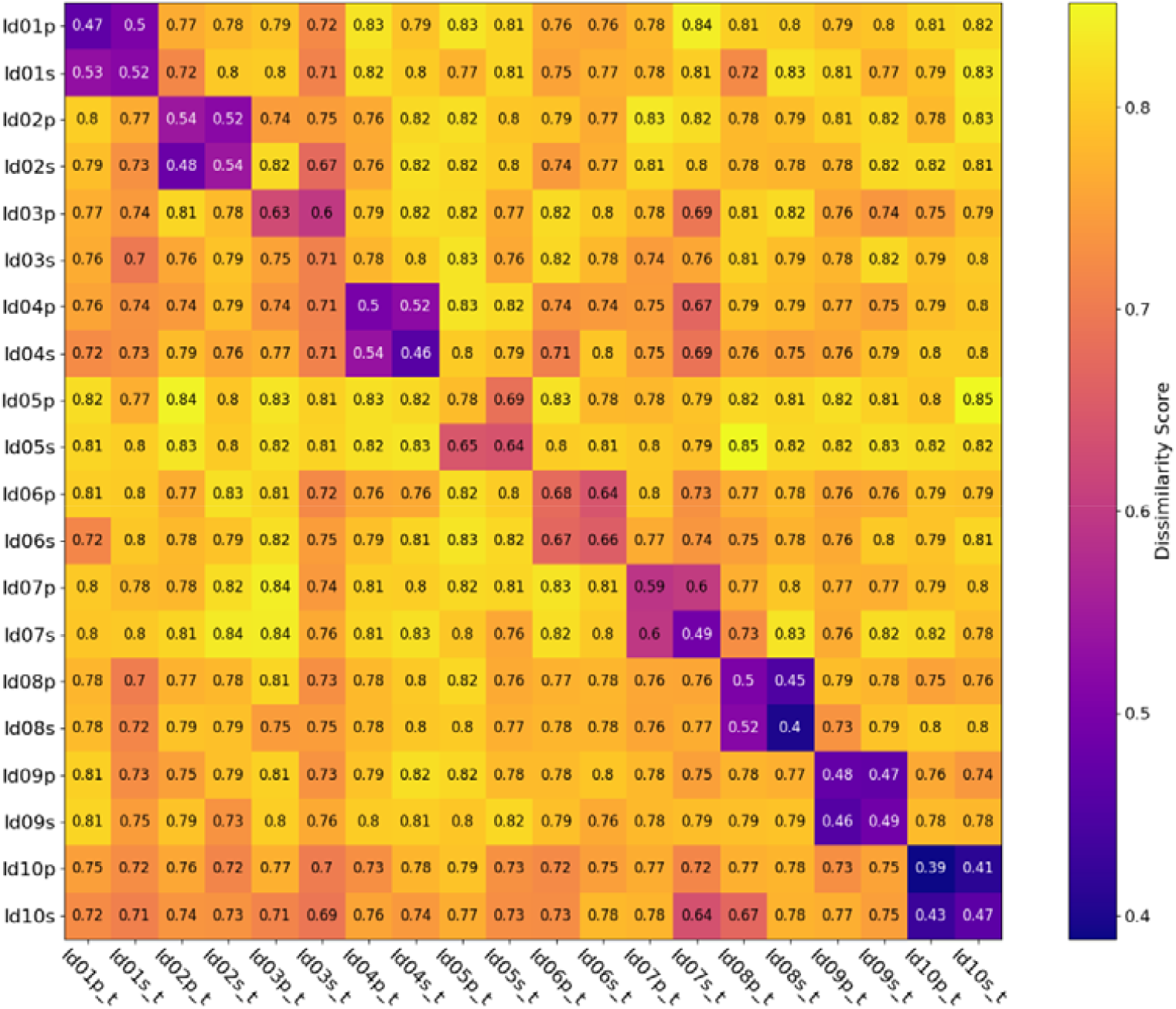
Matrix of dissimilarity scores for each jaw. The id numbers are noted with a p or an s indicating the two scanners and t for post trauma.

A total of 20 dentition surfaces after trauma were quantitively compared to the 20 dentition surfaces prior to trauma, giving 20×20 dissimilarity scores. For each post trauma dentition, there were two comparisons which would constitute a match, one from the same scanner, and one from a different scanner, leaving the remaining 18 comparisons as mismatches.

For 17/20 of the post trauma dentitions, both matches were the lowest scoring comparisons. For the remaining 3/20 (ID03p, ID03s, ID05p), one of the true matches was the lowest scoring comparison, 1/3 (ID03p) being the match from the same scanner. The scores of each comparison can be seen in Figure 7, and the distribution of dissimilarity scores for matches and mismatches can be seen in Figure 8.

**Figure 8.**
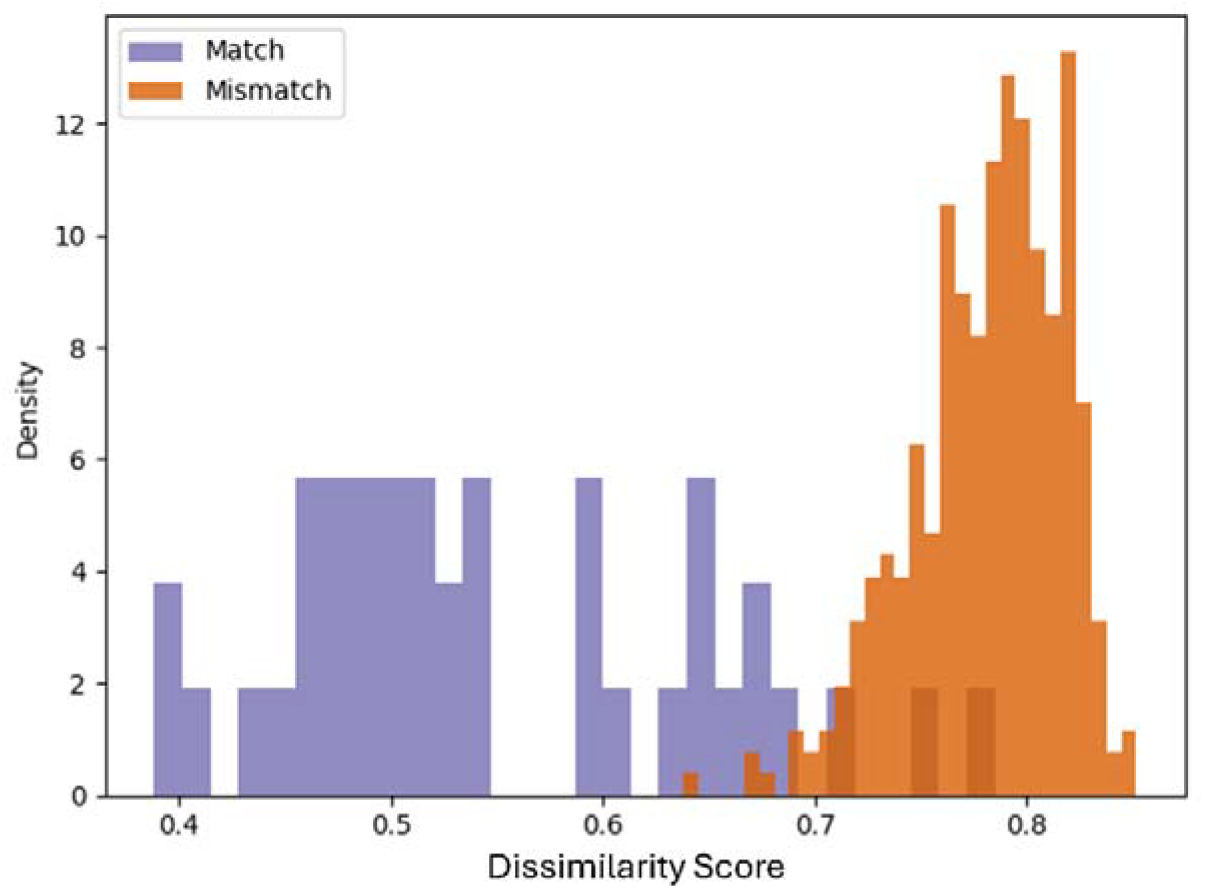
Density distribution of the dissimilarity scores classified by matches and mismatches

Since the keypoint pipeline was branded as a relative scoring scheme [11,14,17], it was not intended to be able to establish an absolute classifier threshold. Nevertheless, a difference between the absolute scores given to a match versus the scores given to a mismatch is seen, with matches scoring on average 0.55 (95% PI 0.40 to 0.75) and mismatches scoring on average 0.78 (95% PI 0.71 to 0.83) (Figure 8).

## DISCUSSION

This study explores the impact of an experimental blunt force trauma on human dentitions. It does so in two parts: one being the descriptions of the dental and bone changes caused by direct blunt force trauma to the approximate occlusal plane to the specimens; the other being the exploration of quantitative dental similarity scoring before and after the blunt force trauma.

In the case of blunt force trauma, this study shows that the curvature signatures on the dentition surface may be preserved enough to show differences between matches and mismatches. Even though this study doesn’t aim at showing an absolute difference in the scores given to matches and mismatches, Figure 7 shows a relative difference in scores, while Figure 8 shows a distinct tendency of difference of absolute dissimilarity scores. This is in line with the intended use of the scoring scheme, as previous publications have underlined its use as an ordering algorithm [11,14,17]. It is hypothesized that a larger difference in dissimilarity score between the lowest and 2^nd^ lowest value will indicate a higher confidence match.

One specimen, ID03, proved more difficult to quantitatively score. This specimen showed both tooth displacement and tooth intrusion, as seen in Figure 6. Nevertheless, at least one of the matching scans for this specimen was scored as the best match, proving that even in such difficult cases, the keypoint pipeline can be of use.

This study is to be seen as an extension of the previous publications regarding the keypoint pipeline [11,13,14,17]. On its own, this study suffers from a limited sample size, as it only includes 10 jaws, and it suffers from a lack of diversity, as it only investigates one type of standardised trauma. Favorably, the specimens comprised a sample of jaws holding a random number of teeth with varying amount and type of dental work, which can be seen as a strength – mimicking a real-life scenario. Though, the results indicate that these parameters, besides jaw type, influenced the resulting types of traumas to both bone and dental structures. But this study is an essential puzzle piece that adds to the confidence in the keypoint pipeline as a tool for the diverse landscape of disaster victim identification.

In regard to trauma, this study lacks variation of the impact, such as variations in trauma direction and inclusion of protective tissue that would be seen in a real-life disaster case [14,19,20]. But to investigate a trauma scenario, standardised trauma allows for systematic investigations of the effects of the trauma, limiting the degrees of freedom. Therefore, this study is limited to investigating the effect of direct blunt force impact on the visible dental surfaces. For future larger trauma studies, more real-life-like scenarios could be established.

The descriptions of the observed tissue destructions in this study add to the understanding of dental surface changes in disaster scenarios featuring blunt force trauma. From Figures 2–6, it is seen that there are extensive changes to the dentition surface, as described in the results section.

Whether the individual curvature signatures on the teeth are preserved well enough for quantitative dental comparison, despite trauma, has not previously been investigated, to our knowledge. Apparently, such an investigation has been limited by the lack of quantitative dental similarity measures that are tooth-position independent. But, with the suggested keypoint pipeline, it is now possible to score curvature similarities of the dentition surface despite displacement [11,14,17]. For such a similarity measure to be proven suitable for the vast diversity of disasters, its performance must be evaluated in a variety of trauma scenarios, including blunt force trauma. The keypoint pipeline has previously been tested in the case of partial jaws, single teeth, and in heat trauma scenarios [11,14,17]. This study extends this list of scenarios where the keypoint pipeline has proven useful.

In conclusion, this study shows that the keypoint pipeline and scoring scheme is able to distinguish 3D dentition surfaces from matching identities and mismatching identities, even in the case of substantial blunt force trauma. It can do so to such an extent that indicates that the pipeline can be a positive addition to the forensic odontology process regarding disaster victim identification. Further studies, focusing on diverse and more complex trauma patterns are suggested as an obvious next step, before the method can be judged as acceptable for application is a real disaster scenario.

## Data Availability

The data of this study can be considered personal data, which the authors are not authorized to share.

## Acknowledgements

We thank Associate Professor Peter Johansen, Department of Electrical and Computer Engineering – Biomedical Engineering, Aarhus University, for access to the high-speed camera for this study. We thank AUFF NOVA for funding this study (Grant number: AUFF-E-2021-9-14).

