## Supplementary Material 1 for "Curvature-based 3D dental comparison to identify trauma-induced surface changes in human teeth: A forensic comparison study" (from another version of this preprint: version 1, 2025-06-20)

**Investigation of Mechanical Trauma on Dentitions: Protocol for Forensic and Experimental Study**

**Study Overview**

This research aims to systematically investigate the similarity of human dentitions before and after mechanical blunt force trauma. By simulating mechanical blunt force trauma in a controlled environment and analysing 3D intraoral dental scans using previously developed similarity measures, the protocol seeks to explore identification options for dentitions exposed to mechanical trauma.

**Study Objective**

1. To expose human dentitions to blunt force trauma and describe the effects
2. To compare dental surfaces from intraoral 3D scans of dentition before and after mechanical blunt force trauma
3. To investigate blunt force traumas effect on the identifiability of dentitions by 3D surface scans

**Study Design**

This study will consist of two parts:

1. **Visual Forensic Odontology Analysis**: Examination and description of dentitions before and after dental trauma.
2. **3D dental surface comparison**: Comparison of dental surface similarity before and after dental trauma.

**Study Population**

Anonymous human dentition samples (partial jaws with teeth) from preserved dental specimens will be used for mechanical trauma testing.

The power calculation below is based on distribution of pooled similarity scores of dentitions in other scenarios (heat trauma, 6 months’ time difference, single teeth vs full jaws).
To clarify if there is a difference in scores between scans originating from the same individual (matches) and scans originating from different individuals (mismatches), the score distributions will be subject to a Mann-Whitney U test.

$$Effect size \left( \Delta\right)\left( Rank-biserial correlation \right)=0.966 -{0.2}^{*}$$

$$* =Correction due to expected smaller effect size$$

$$Significance level \left( \alpha\right)=0.01$$

$$\mathrm{Power}\left( 1-\beta\right)=0.9$$

$$Minimum sample size per group \left( n \right)=6$$

$$Number of groups (k)=2$$

Power calculations were done through Monte Carlo simulation

**Inclusion Criteria for Experimental Samples**

Jaws with teeth from human adult individuals.

**Exclusion Criteria for Experimental Samples**

Pices smaller than 5x5 mm after trauma exposure

**Materials and Methods**

**1. Data Collection – Visual Forensic Analysis**

- Detailed forensic odontology description of the dental surfaces of the jaw part and teeth prior to trauma
- 3D intraoral scan of dental surfaces prior to trauma
- Detailed forensic odontology description of the dental surfaces of the jaw part and teeth after trauma
- 3D intraoral scan of dental surfaces after trauma

**2. Experimental Simulation – Mechanical Trauma Testing setup**

The following setup was inspired by *Houg, K.P., Adanty, K., MacGillivray, S.R., McAllister, L., Levin, L., Alexiou, M., Graf, D., Romanyk, D.L. and Dennison, C.R. (2021), On the ability of experimental impact measures to predict tooth injuries in an ex vivo swine model. Dental Traumatology, 37: 464-473.* [*https://doi.org/10.1111/edt.12645*](https://doi.org/10.1111/edt.12645)

- **Physical setup**
  - A) Pipe (2 m, 10 cm Ø), B) Flat-bottomed flask of sturdy material (2000 ml, <10 cm Ø), C) High-speed camera, D) Textile scarf, needle and thread, E) Tape or the like for fastening A

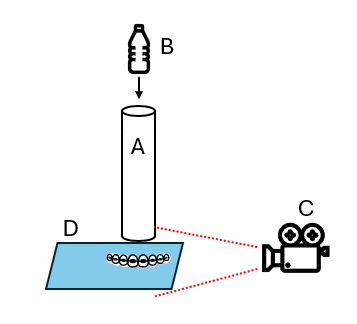

- **Experiment setup - calibration**
  - Fasten the pipe (A) in a vertical position, 10-15 cm above the resting surface of impact
  - Setup the high-speed camera to capture at least the 10-15 cm between the resting surface of impact and the pipe
  - Fill the bottle (B) with the material of choice (water+sand)
  - Weigh the bottle
  - Make calibration experiments by dropping the bottle from the pipe entrance and ensure that the high-speed camera captures the area of interest.
  - Repeat this setup at least 3 times.

Estimated impact (F) measured in Newton (kg*m/s^2^) can be calculated as follows:

$$Collision speed \left( v1 \right)=\sqrt{2*g*H},$$

$$g=\frac{9.816m}{s^{2}},$$

$$H=distance from pipe entrance to resting surface \left( \mathrm{meters} \right),$$

$$F=\frac{M*\left( v1-v2 \right)}{t},$$

$$M=mass of bottle \left( kilograms, kg \right),$$

$$Time of impact \left( t \right)=\frac{n}{\mathrm{fps}},$$

$$n=number of frames in which there is contact between the bottle and the resting surface,$$

$fps=frames per second of the high speed camera (for precision)$,

$$Speed after collision \left( v2 \right)=e*v1,$$

$$Restitution coefficient \left( e \right)=\sqrt{\frac{h}{H}} ,$$

$$h=rebound height of the bottle after colliding with the resting surface$$

Final equation will therefore be:

$$F=\frac{M*\left( \sqrt{2*\frac{9.816m}{s^{2}}*H}-\left( \sqrt{\frac{h}{H}}*\sqrt{2*\frac{9.816m}{s^{2}}*H} \right) \right)}{\frac{n}{\mathrm{fps}}}$$

F being the Force,
M being the mass of the falling object,
h being the rebound height of the falling object,
H being the distance the object falls,
n being the number of frames where there is contact,
fps being the frames per second of the high-speed camera.

- **Experiment setup – measures**

M being the mass of the falling object	2 kg
h being the rebound height of the falling object	0.12 m
H being the distance the object falls	2 m
n being the number of frames where there is contact	6
fps being the frames per second of the high-speed camera	2000 1/s

F	~ 3154 N

- **Experiment setup – execution**
  - Fasten the jaw to the fabric using needle and thread (D)
  - Ensure the high-speed camera is in place
  - Place the jaw and fabric below the exit of the pipe (A)
  - The bottle (B) is dropped from the entrance of the pipe
  - The estimated impact can be corrected based on the time of impact and the speed after collision as described during calibration.

1. **Data Analysis**

- **Visual Forensic Analysis**:
  - The trauma effect on the dentition surface is described in detail by a forensic odontologist.
- **3D dental surface comparison**:
  - The 3D intraoral scans of the dentition surfaces are subject to the developed keypoint pipeline to assess the quantitative surface similarity before and after trauma.

**Ethical Considerations**

- **Approval**: Ethical approval for experimental studies using human dental samples have been obtained from the relevant institutions.
- **Safety**: All procedures involving mechanical trauma simulation will adhere to safety standards to eliminate risk to researchers.

**Timeline**

- **Phase 1**: Physical sample collection and acquisition of experimental setup (1 month)
- **Phase 2**: Experimental testing (1 week)
- **Phase 3**: Data analysis and interpretation (1 months)
- **Phase 4**: Reporting results in scientific paper format (1 month)
- **Total Duration**: approximately 4 months

**Budget**

The estimated budget for this project will include:

- Experimental equipment: pipe, bottle, high-speed camera, textile scarfs, needles and threat, duct tape,
- Human dentition samples; available in the research group
- Software for data analysis; in-house custom software
